# Artificial Intelligence, LLM-based generation of synthetic patients with Parkinson’s Disease: towards a digital twin paradigm for in silico studies

**DOI:** 10.64898/2026.04.28.26351471

**Authors:** Emilio Merlo Pich, Octavio Pomponio, Maria Antonietta Magno, Michele Berti, Lorenzo Li Lu, Andrea Coser, Ilaria Cani, Giovanna Calandra Buonaura, Manfredi Valenti, Camilla Cividini, Nicoletta Semenzato, Anna Baggi, Sebastiano Saccani, Jean Marie Franzini

## Abstract

Heterogeneity in sporadic Parkinson’s Disease (PD) is a critical problem that drives variable rates of progression and treatment response and complicates clinical trials. Access to large PD datasets that may help in clustering this heterogeneity is restricted by privacy and regulatory constraints. Simulated patients or digital twins may offer a solution. We developed a large language model (LLM)-framework to generate high-fidelity synthetic PD patients from the Parkinson’s Progression Markers Initiative (PPMI) dataset based on the open-source Qwen3-8B-Base model. Using a relational, tree-structured representation and domain-specific fine-tuning, the model produces patient-level records with longitudinal clinical, imaging, and biomarker data. Fidelity was assessed through distributional similarity, correlation structure, and neurologist review. Utility was tested by training diagnostic classifiers, reproducing a published pharmacometric disease progression model applied to in silico trials, and by extracting a stringent dopamine–motor impairment relationship at early PD stages. Privacy was evaluated via identical match share, distance-to-closest-record, and membership inference attacks. These findings support the use of a dedicated LLM framework for patient simulation, contributing to the foundations of digital twins for PD in silico trials.

## 1 Introduction

Parkinson’s disease (PD) is the second most common neurodegenerative disorder worldwide and affects 7–10 million individuals, imposing a substantial clinical and socioeconomic burden [1]. Its core pathophysiology is characterized by progressive degeneration of dopaminergic neurons in the substantia nigra pars compacta, leading to dopamine depletion along nigrostriata pathways to caudate and cardinal motor symptoms including bradykinesia, rigidity, tremor, and postural instability [2, 3]. Beyond motor signs, PD encompasses a wide spectrum of non-motor manifestations, such as sleep and autonomic dysfunction, mood and cognitive disturbances and sensory complaints, that strongly impact quality of life and care needs [3]. Motor impairments are generally anticipated by prodromal symptoms, affecting olfaction, sleep, gastrointestinal system or mood changes [4].

Despite decades of research, no disease-modifying treatment has yet been approved. Levodopa and other dopaminergic therapies provide symptomatic benefit but are associated with motor fluctuations and dyskinesias as the disease progresses. A central challenge for both clinical management and drug development is the pronounced inter-individual heterogeneity in disease presentation, progression rate, and treatment response. Patients with similar baseline diagnoses can follow divergent trajectories, from rapidly progressive forms to slowly evolving phenotypes with prolonged functional independence [5, 6]. Longitudinal analyses of large cohorts have identified distinct progression subtypes, including motor-dominant, cognitive-dominant, and mixed profiles [7], suggesting underlying biological diversity in pathophysiological mechanisms and reserve, that could be described within a ‘biological staging’ framework [8].

The Parkinson’s Progression Markers Initiative (PPMI) [9] is one of the most comprehensive longitudinal observational cohorts in PD, with standardized serial assessments across clinical scales, dopamine transporter imaging (DScan), cerebrospinal fluid and blood biomarkers, as well as genetic measures in de novo PD, prodromal individuals, and controls. PPMI and related efforts have enabled data-driven phenotyping, prognostic model development, and mechanistic modeling of PD progression due to the informed consent signed at the time of data consenting [10, 11]. However, considering other databases, broad reuse of data from is limited by privacy regulations (e.g., GDPR), legacy consent forms, and limiting data-sharing agreements that constrain cross-institutional and cross-border access to patient-level records. These constraints hinder replication, external validation, and model-based trial design [12, 13].

Synthetic data, virtual patients and digital twins have emerged as promising solutions to these barriers by generating artificial records that preserve statistical and clinical structure without corresponding to identifiable individuals. In parallel, the concept of digital twins (patient-specific computational models that integrate clinical, imaging, and biological information to simulate disease evolution and treatment effects) is rapidly expanding in cardiovascular disease, oncology, and neurology [14–16]. In PD, mechanistic models linking dopaminergic degeneration to motor deficits and large-scale network dynamics have been used to investigate disease progression and treatment effects, including dopaminergic replacement and deep brain stimulation [17–21].

Recent advances in large language models (LLMs) extend synthetic data generation beyond traditional generative models by exploiting rich prior knowledge and structured generation capabilities [22–25]. Recent text-to-tabular approaches have demonstrated that LLMs can produce realistic neurological patient-level records across multiple domains, provided that domain-specific constraints and privacy evaluations are rigorously applied [24, 25]. Yet, most existing work focuses on cross-sectional data, single tables, or generic benchmarks and does not address mechanistic time-dependent relationships crucial for disease modeling and regulatory-grade in silico trial simulations.

Here we build on PPMI and an LLM-based synthetic data framework originally developed on an open-source Qwen3 architecture [26, 27] to generate longitudinal synthetic PD patients structured as relational trees with demographics, repeated clinical visits, imaging and biospecimens. We ask whether fine-tuned LLMs can produce synthetic cohorts that (i) match real data distributions and correlation structures, (ii) preserve mechanistic links between dopaminergic imaging and motor impairment, and (iii) confirm published results obtained with longitudinal disease progression modeling on real PD patient data, while (iv) satisfying stringent privacy requirements. Our objective was to demonstrate a concrete, quantifiable bridge from real patient statistics to healthcare digital twins suitable for in silico trial design and future regulatory applications.

## 2 Results

### 2.1 Cohort and synthetic population overview

We extracted a longitudinal dataset from PPMI as of June 2024, including PD, prodromal, and healthy control participants with multiple years of follow-up and low missingness across 92 clinically, imaging, and biologically relevant variables (see Table 4.1 in Section 4.1). Data encompassed, among others, demographics, Movement Disorder Society–Unified Parkinson’s Disease Rating Scale (MDS-UPDRS) components (including part III motor items OFF and ON medication), Hoehn and Yahr staging, cognitive scales, DaTSCAN measures, and cerebrospinal fluid (CSF) biomarkers. The relational structure comprised a root patient table and two child tables for longitudinal clinical assessments and biospecimens. Thirty-two files were included, with a total number of subjects described in Table 2.1

**Table 2.1:** Cohort composition of the PPMI dataset used in this study.

| COHORT_DEFINITION | Description | number of patients |
| --- | --- | --- |
| Parkinson’s Disease (PD) | People who have a formal diagnosis of PD. | 1870 |
| Prodromal (PR) | People who are at risk of developing PD based on clinical features, genetic variants or other biomarkers but have not been formally diagnosed. | 1988 |
| Healthy Control (HC) | People with no neurologic disorder and no first-degree relative with PD. | 393 |
| SWEDD | Participants who were clinically diagnosed with PD but had a normal DAT SPECT. | 81 |
| All cohorts |  | 4332 |

Using this dataset, we trained and evaluated an LLM-based synthetic data generator, running on a Qwen3-8B-Base machine, that outputs patient-level JSON objects encoding the full relational tree. The synthetic population matched the original cohort in size for primary analyses, with additional oversampled variants used for specific mechanistic evaluations.

### 2.2 Fidelity of key parameter univariate marginal distributions

We first assessed univariate fidelity by comparing baseline distributions of key demographic and clinical variables between real and synthetic datasets. Fig. 2.1 shows representative variables including enrollment age (`ENROLL_AGE`), OFF-state MDS-UPDRS part III total score (`NP3TOT_OFF`), and the key molecular biomarker (`CSF Alpha-synuclein`). Real and synthetic distributions largely overlapped, with comparable central tendency, dispersion, and tail behavior.

**Figure 2.1:**
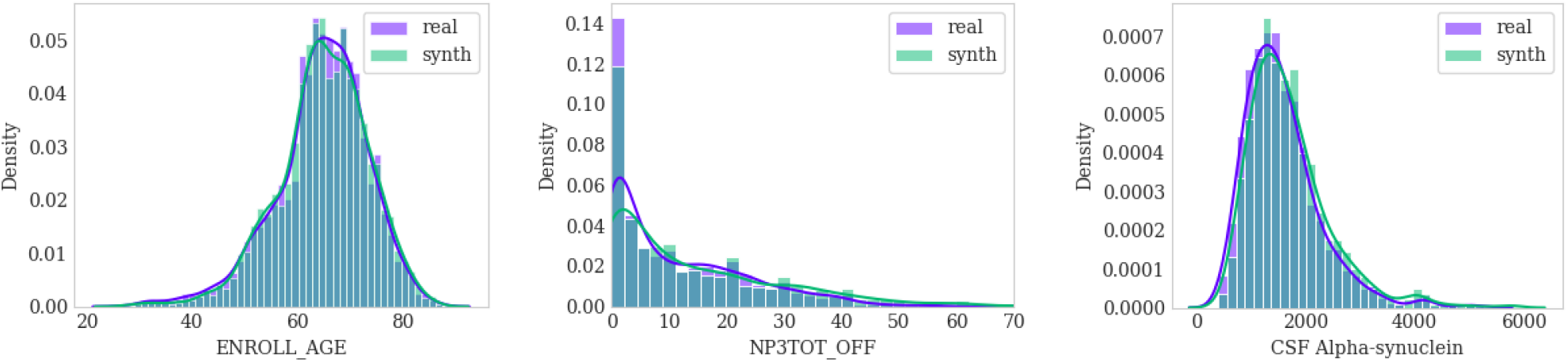
Univariate marginal distributions for the static variable `ENROLL_AGE` and the three variables `NP3TOT_OFF` and `CSF Alpha-synuclein` computed at the baseline. The plots compare real (*purple*) and synthetic (*green*) distributions, with histograms and kernel density estimation (KDE) curves (in solid lines) showing the smooth probability density functions.

Quantitatively, we applied several complementary metrics (see Table 2.2). For continuous variables, the Kolmogorov–Smirnov (KS) test was often formally significant, reflecting its sensitivity in large samples to small local discrepancies. However, Jensen–Shannon divergence values were low, indicating information-theoretic similarity between distributions. For categorical variables, Kullback–Leibler divergence values were small across diagnostic groups and sex, consistent with preserved category proportions.

**Table 2.2:**
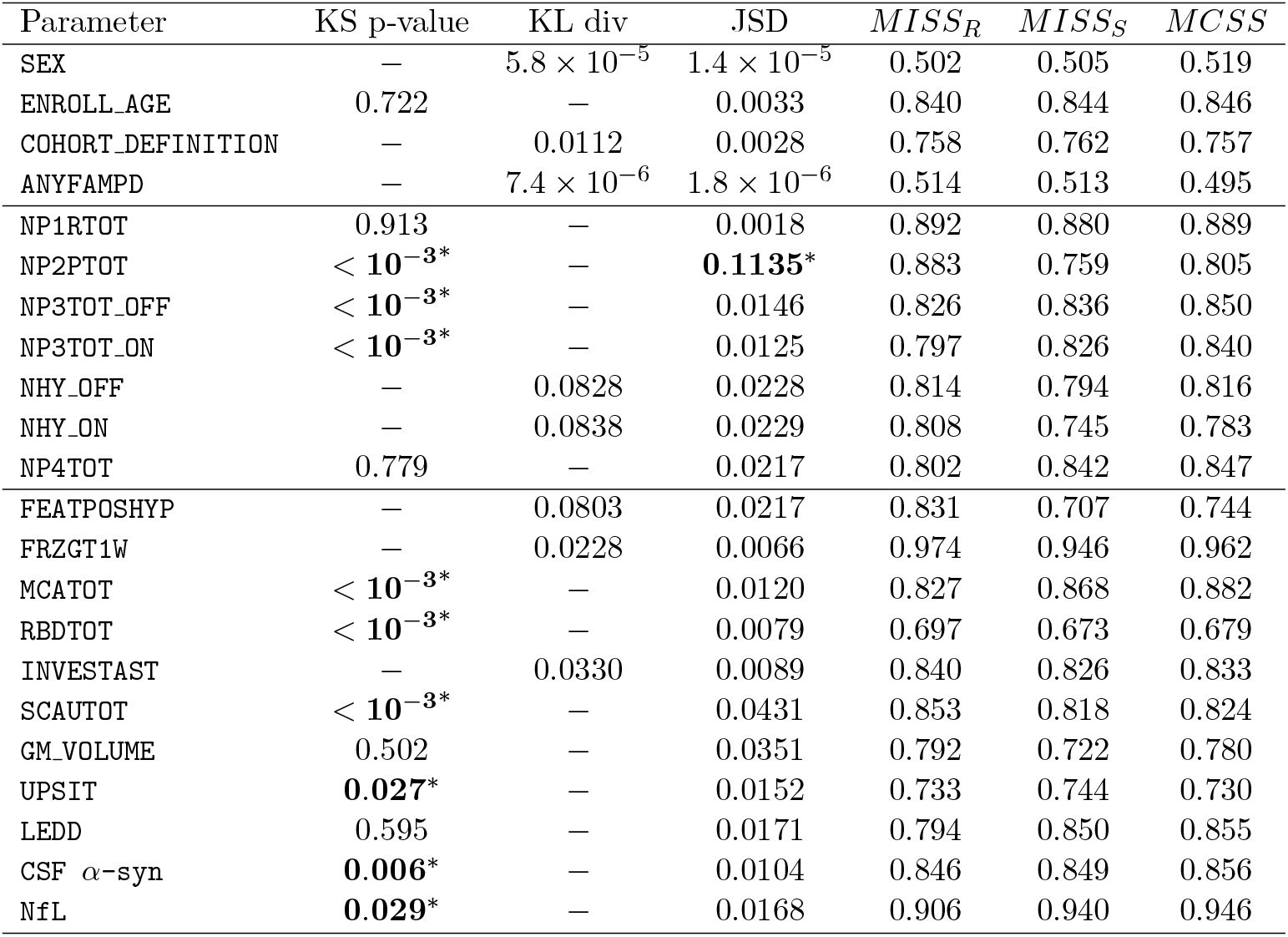
Comparison of metrics described in Section 2.2 between real and synthetic data. An asterisk (*) indicates that the statistical test was not passed. For simplicity, NP3 subcomponents are omitted from the table.

| Parameter | KS p-value | KL div | JSD | $MISS_R$ | $MISS_S$ | $MCSS$ |
| --- | --- | --- | --- | --- | --- | --- |
| SEX | — | $5.8 \times 10^{-5}$ | $1.4 \times 10^{-5}$ | 0.502 | 0.505 | 0.519 |
| ENROLL_AGE | 0.722 | — | 0.0033 | 0.840 | 0.844 | 0.846 |
| COHORT_DEFINITION | — | 0.0112 | 0.0028 | 0.758 | 0.762 | 0.757 |
| ANYFAMPD | — | $7.4 \times 10^{-6}$ | $1.8 \times 10^{-6}$ | 0.514 | 0.513 | 0.495 |
| NP1RTOT | 0.913 | — | 0.0018 | 0.892 | 0.880 | 0.889 |
| NP2PTOT | $< 10^{-3*}$ | — | **0.1135\*** | 0.883 | 0.759 | 0.805 |
| NP3TOT_OFF | $< 10^{-3*}$ | — | 0.0146 | 0.826 | 0.836 | 0.850 |
| NP3TOT_ON | $< 10^{-3*}$ | — | 0.0125 | 0.797 | 0.826 | 0.840 |
| NHY_OFF | — | 0.0828 | 0.0228 | 0.814 | 0.794 | 0.816 |
| NHY_ON | — | 0.0838 | 0.0229 | 0.808 | 0.745 | 0.783 |
| NP4TOT | 0.779 | — | 0.0217 | 0.802 | 0.842 | 0.847 |
| FEATPOSHYP | — | 0.0803 | 0.0217 | 0.831 | 0.707 | 0.744 |
| FRZGT1W | — | 0.0228 | 0.0066 | 0.974 | 0.946 | 0.962 |
| MCATOT | $< 10^{-3*}$ | — | 0.0120 | 0.827 | 0.868 | 0.882 |
| RBDTOT | $< 10^{-3*}$ | — | 0.0079 | 0.697 | 0.673 | 0.679 |
| INVESTAST | — | 0.0330 | 0.0089 | 0.840 | 0.826 | 0.833 |
| SCAUTOT | $< 10^{-3*}$ | — | 0.0431 | 0.853 | 0.818 | 0.824 |
| GM_VOLUME | 0.502 | — | 0.0351 | 0.792 | 0.722 | 0.780 |
| UPSIT | **0.027\*** | — | 0.0152 | 0.733 | 0.744 | 0.730 |
| LEDD | 0.595 | — | 0.0171 | 0.794 | 0.850 | 0.855 |
| CSF α-syn | **0.006\*** | — | 0.0104 | 0.846 | 0.849 | 0.856 |
| NfL | **0.029\*** | — | 0.0168 | 0.906 | 0.940 | 0.946 |

To capture similarity in mixed-type clinical profiles, we computed Gower distance–based measures: mean intra-sample similarity for real data (*MISS*_*R*_), for synthetic data (*MISS*_*S*_), and mean cross-sample similarity between real and synthetic records (*MCSS*). *MISS*_*R*_, *MISS*_*S*_ and *MCSS* values were close across variables, indicating that that neighborhood structure is well preserved between real and synthetic datasets. Overall, these results indicate that the synthetic records were on average as similar to real ones as real patients are to each other, without evidence of overfitting or collapse.

### 2.3 Fidelity as preservation of parameter correlation structure and longitudinal dependencies

We next evaluated whether the synthetic cohort preserved multivariate relationships at baseline and over time. At baseline, we computed Pearson correlations among key demographic, motor, non-motor, imaging, and biomarker variables. The correlation heatmaps for real and synthetic data (Fig. 2.2) showed similar patterns with some differences in the strength of correlation, including positive associations among MDS-UPDRS subscales and `SCAUTOT`, with an inverse relationships with alpha-synuclein levels in CSF and with the imaging gray matter brain `GM_VOLUME` that show a strong inverse correlation with `NfL` peripheral markers of neurodegeneration, as expected.

**Figure 2.2:**
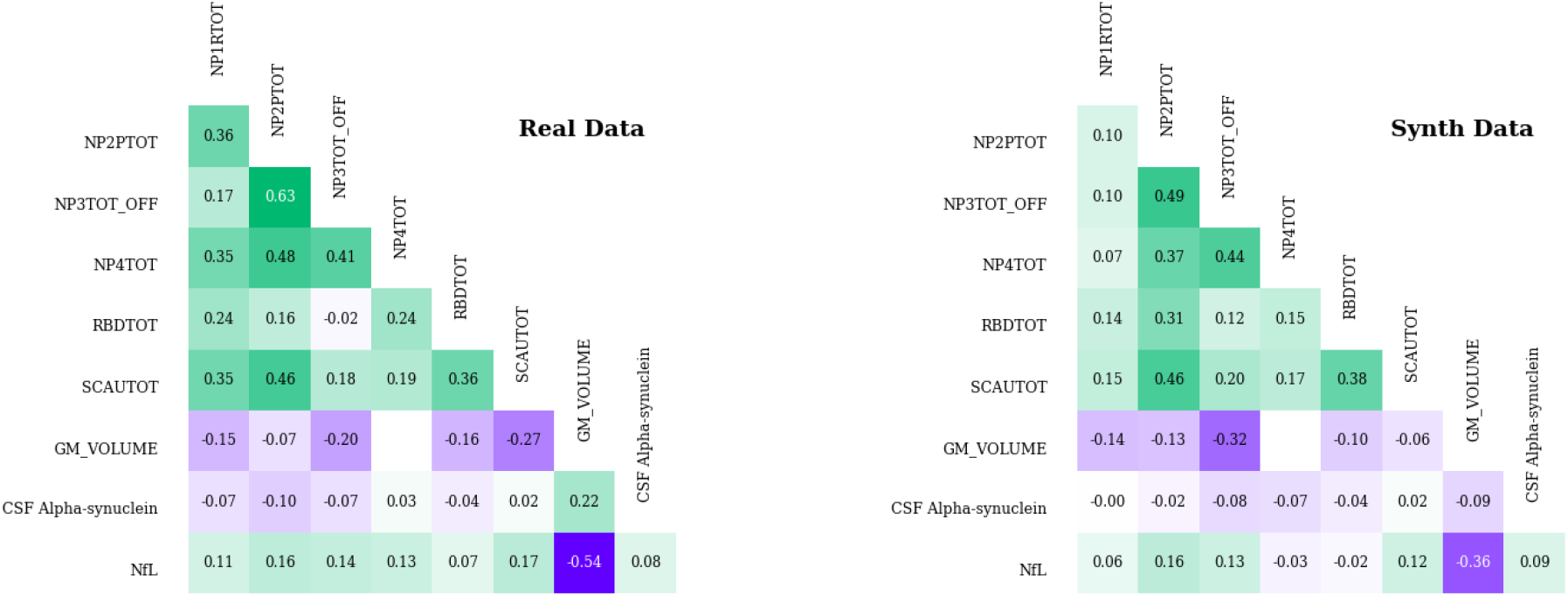
Comparison of Pearson correlations between selected variables of the real and synthetic datasets computed at baseline, illustrating that the synthetic data reproduce the overall correlation patterns observed in the real data, with a high degree of structural similarity.

To account for repeated measurements, we used repeated-measures correlation (rmcorr) to estimate within-subject associations between longitudinal variables. For PD participants, we examined relationships between OFF-state motor scores, non-motor scores, Hoehn and Yahr stage, and selected biomarkers, including the dopaminergic deficit imaging markers `DATSCAN_CAUDATE_R`. The rmcorr matrices for real and synthetic cohorts (Fig. 2.3) were closely aligned, with minor deviations well within confidence intervals. Synthetic data preserved both the direction and approximate magnitude of longitudinal associations, including increasing motor impairment severity co-evolving with age, worsening postural instability and stable or mildly declining cognition. These results indicate that the LLM-based generator captured not only marginal distributions, but also higher-order patterns of covariation critical for disease modeling and subtype characterization.

**Figure 2.3:**
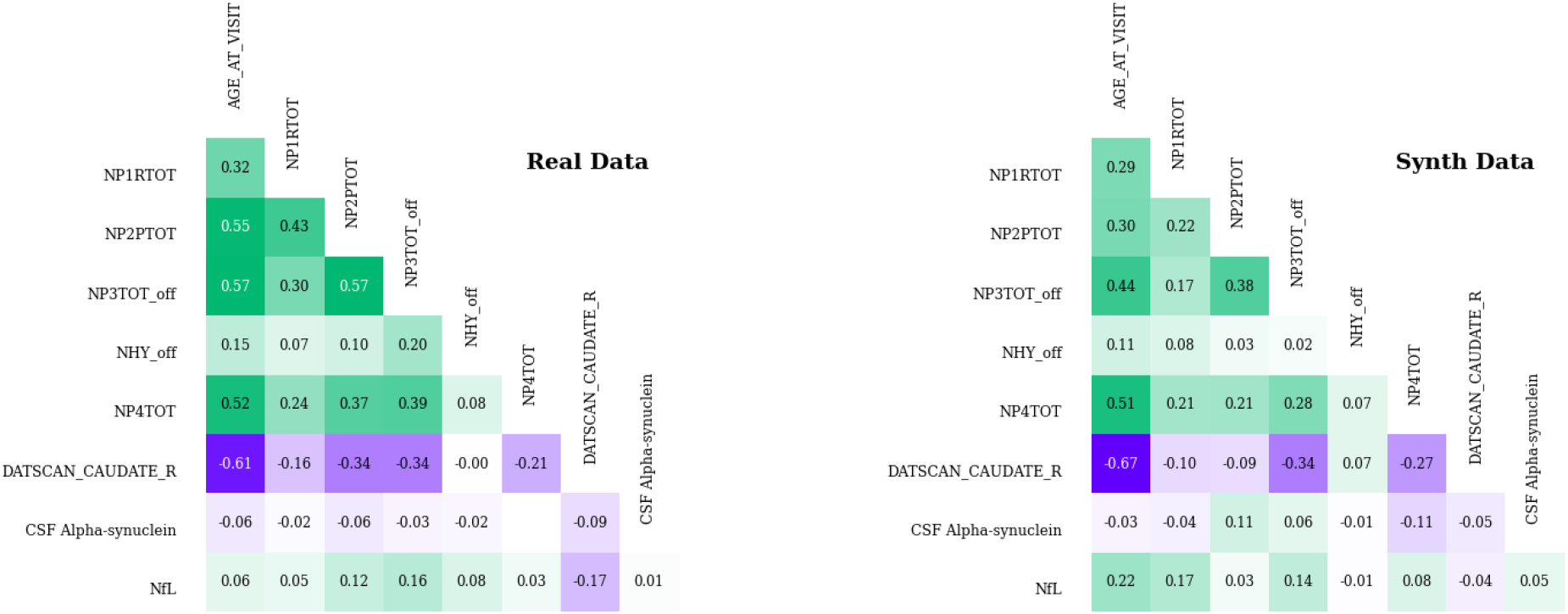
Comparison of repeated measures correlation (rmcorr) matrices between real and synthetic data for PD cohorts. Each heatmap shows the pairwise rmcorr coefficients for longitudinal variables, capturing the within-individual association across multiple time points. The left panel represents the real dataset, while the right panel represents the synthetic dataset. High similarity between the heatmaps indicates that the synthetic data preserves the intra-individual relationships observed in the original data over time.

### 2.4 Clinical plausibility assessment by neurologists

To complement data statistical metrics with domain expertise, we performed a blinded clinical plausibility assessment asking two neurologists to provide diagnostic judgment (PD vs non-PD) and plausibility rating (typical vs implausible PD, or control profile) on patient’s profile “flash cards” containing 36 demographic, clinical, imaging, and biomarker variables for each case. Inter-rater agreement among neurologist on diagnosis was substantial (Cohen’s *κ* = 0.62), with accuracies of 100% and 95% relative to known labels, and no significant difference between real and synthetic cases. Plausibility ratings showed lower concordance (*κ* = 0.09), with an accuracy of 95% and 47%, respectively, reflecting expected individual variability in clinical thresholding by the two differently trained neurologists. Interestingly, again, there was no significant difference in plausibility distribution between real and synthetic cohorts when tested via Fisher’s exact test. Synthetic profiles were therefore indistinguishable from real cases at the level of expert qualitative assessment.

### 2.5 Predictive utility: diagnostic classification

We evaluated task-specific utility by training a multiclass classifier to distinguish PD, prodromal, and healthy control participants using only baseline demographics and clinical scores, mimicking a real-world diagnostic setting. A Random Forest model was trained either on real data and tested on held-out real data (Real → Real) or trained on synthetic data and tested on the same held-out real set (Synth → Real).

Fig. 2.4 shows ROC curves for both settings. When trained and tested on real data, the model achieved high discriminative performance for all three classes, with AUC values consistent with prior work on real patient PPMI-based diagnostic models [11,18,20]. When trained on synthetic data, AUCs remained closely aligned with the Real → Real benchmark, including for the challenging prodromal vs control discrimination. Differences in AUC and calibration were small and within expected variability for resampling-based evaluations.

**Figure 2.4:**
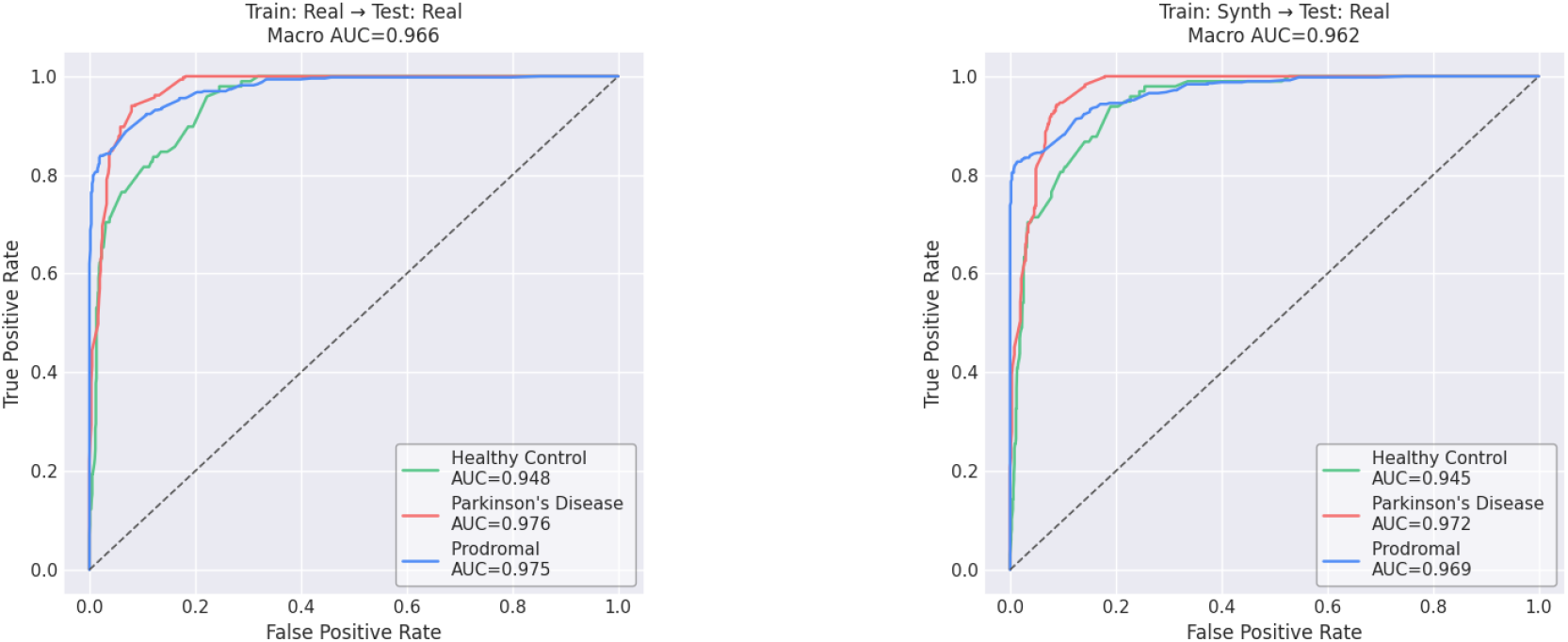
Multi-class ROC curves for the Random Forest classifier trained either on real data (*left panel*) or on synthetic data (*right panel*) and evaluated on real patients. All models were trained using only baseline clinical assessments and demographic variables, reflecting a real-world diagnostic setting in which predictions must be made at the first clinical visit. An identical learning pipeline was used in both cases, consisting of one-hot encoding and a Random Forest classifier trained on a stratified 75%*/*25% split. Class-specific AUCs were highly consistent between the Real → Real and Synth → Real scenarios, indicating minimal performance degradation. The close alignment of the ROC curves confirms that synthetic data faithfully reproduce the discriminative structure present in the real dataset, supporting real/synthetic interchangeability for baseline diagnostic modeling.

These findings demonstrate that classifiers trained solely on LLM-generated synthetic data generalize effectively to real patients, indicating that the synthetic cohort preserves discriminative structure and supports development of diagnostic algorithms.

### 2.6 Analytical utility: reproducing a pharmacometric PD progression model

To test whether synthetic data support mechanistic modeling, we reproduced the nonlinear mixed-effects disease progression model proposed by Ribba et al. (2024) [20], which describes OFF-state MDS-UPDRS part III trajectories under symptomatic pharmacological treatment. The model combines logistic growth for natural disease progression with a treatment effect driven by pharmacological treatments expressed as levodopa equivalent daily dose (LEDD) and quantifies inter-individual variability in baseline severity, progression rate, plateau, and drug response.

Using the nlmixr2 framework, we fitted the model separately to real and synthetic datasets, using identical inclusion criteria and preprocessing. Parameter estimation employed the SAEM algorithm with log-normal distributions for structural parameters and normal distribution for the treatment effect. Population-level parameter estimates for baseline motor score, progression rate, and treatment effect overlapped between real and synthetic cohorts within 95% confidence intervals, as showed in Fig. 2.5, left panel. To be noted, the plateau parameter (*θ*) was higher in synthetic data, indicating modest overestimation of long-term disease severity; however, the mean ±1 SD ranges of the two distributions overlap.

**Figure 2.5:**
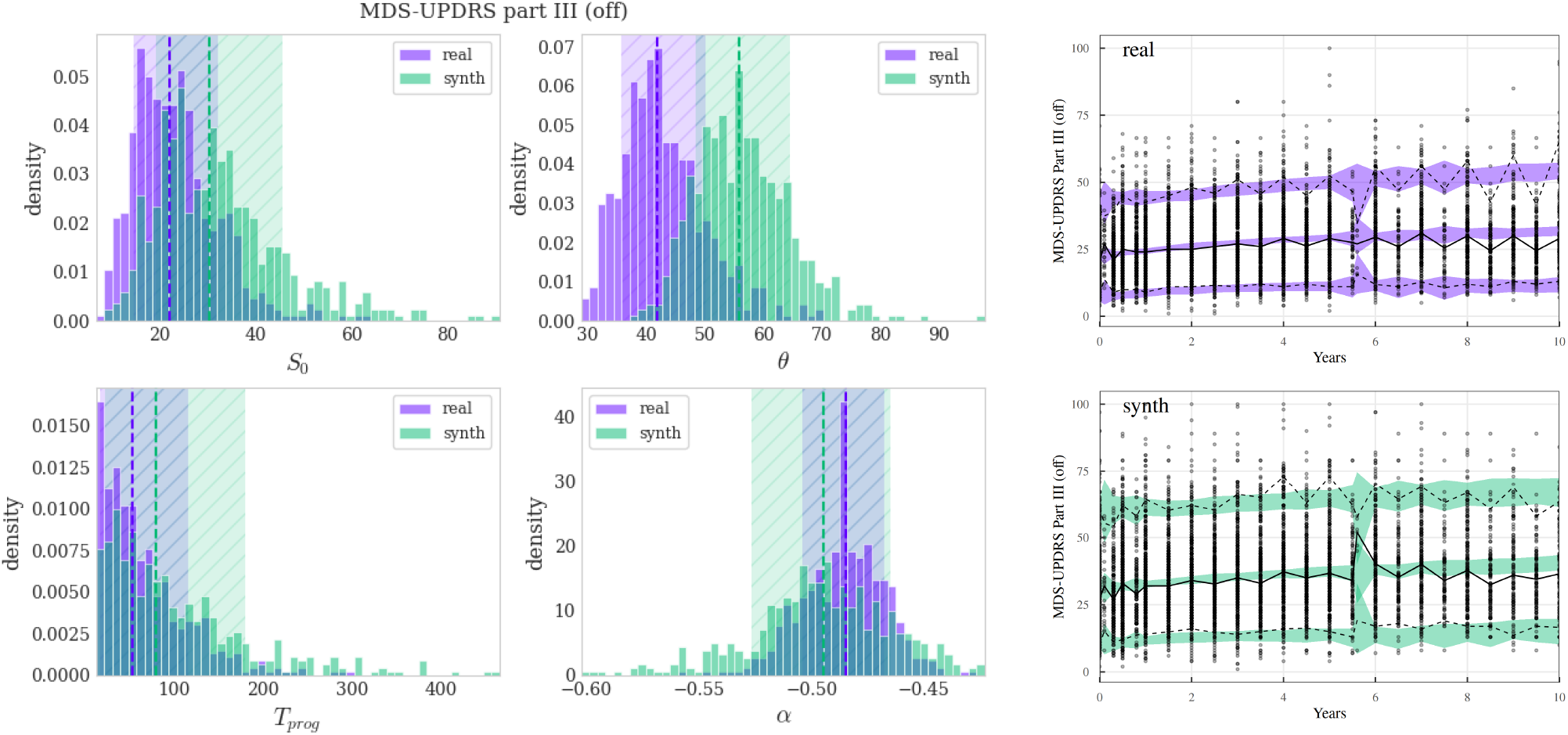
*Left panel* - Distributions of the estimated model parameters for both real (*purple*) and synthetic (*green*) datasets. The dashed vertical lines indicate the median of each parameter distribution, while the shaded regions represent the range of one standard deviation around the mean. *Right panel* - Visual Predictive Check comparing model predictions to observations for real (top) and synthetic (bottom) datasets showing MDS-UPDRS Part III scores over 10 years. Shaded bands indicate 90% prediction intervals from 100 simulations. Black lines show observed 5th, 50th (solid), and 95th percentiles. Points represent individual observations. Both datasets show good agreement between model predictions and observations.

### 2.7 Mechanistic utility: dopamine-motor relationship

A central requirement for developing digital twins from a simpler patient simulation is the faithful representation of key mechanistic relationships between symptoms and the underlying pathology [14]. Experimental lesion models in animals and human clinical-imaging and post-mortem studies in PD patients have long shown that reduced striatal dopaminergic terminals correlate with bradykinesia and akinesia severity, considered as one hallmark of disease [11, 28, 29].

To support this relationship also in the synthetic cohort, we analyzed the joint longitudinal relationship between OFF-state MDS-UPDRS part III scores (`NP3TOT_OFF`) and DaTSCAN signal in the left caudate nucleus (`DATSCAN_CAUDATE_L`) using linear mixed models and repeated-measures correlation, decomposing the bivariate correlation into within-subject and between-subject components. The data of the first 4 years since diagnosis of three populations were considered: real data (*n* = 329), synthetic data (*n* = 135), and oversampled synthetic data (*n* = 647). Estimated model parameters for the 3 populations are reported in Table 2.3.

**Table 2.3:** Bivariate longitudinal analysis across datasets. *β*_*X*_ and *β*_*Y*_ are population slopes (units/year) from LMM. *ρ*_within_ is the repeated-measures correlation [30] with analytic 95% CI. *ρ*_between_ is the correlation of random intercepts with bootstrap 95% CI (1000 resamples).

| Dataset | $\beta_X$ | $\beta_Y$ | $\rho_{\text{within}}$ [95% CI] | $p$ | $\rho_{\text{between}}$ [95% CI] | $p$ |
| --- | --- | --- | --- | --- | --- | --- |
| Real | +2.470 | -0.120 | -0.370 [-0.430, -0.310] | $2.1 \times 10^{-28}$ | -0.175 [-0.279, -0.073] | $1.4 \times 10^{-3}$ |
| Synth | +2.703 | -0.134 | -0.311 [-0.410, -0.210] | $1.7 \times 10^{-8}$ | -0.111 [-0.278, +0.059] | $2.0 \times 10^{-1}$ |
| Synth_over | +2.926 | -0.137 | -0.374 [-0.420, -0.330] | $6.8 \times 10^{-52}$ | -0.126 [-0.200, -0.047] | $1.4 \times 10^{-3}$ |

In real data, OFF-state motor severity increased by approximately 2.5 points per year, while DaT signal declined by ∼ 0.12 units per year, with significant negative within-subject correlation (*ρ*_within_ ≈ −0.37) and between-subject correlation (*ρ*_between_ ≈ −0.18). Synthetic data, and especially those of the oversampled synthetic cohort, reproduced these slopes and correlations within confidence intervals, with a less stringent results shown by the smaller size synthetic cohort. This preservation of a quantifiable dopaminergic–motor coupling establishes that LLM-based synthetic patients capture a mechanistic axis central to PD pathophysiology and staging. It moves beyond generic distributional similarity and supports the interpretation of synthetic records as pathophysiologically grounded simulated patients, potentially suitable for digital twin instantiation in mechanistic models of basal ganglia and cortical network dynamics. However, since other pathophysiological mechanistic and symptoms are present during PD progression, more work is needed before establishing a proper digital twin for PD patients that would reflect a recent staging proposals that integrate clinical, imaging and biomarkers into a unified framework [8].

### 2.8 Privacy evaluation

Ensuring that synthetic data do not compromise participant privacy is essential for regulatory acceptance and broad sharing. We implemented a three-layer privacy evaluation framework grounded in recent recommendations for health data synthesis [31, 32].

First, identical match share (IMS) quantified direct copying of records. The value of *f*_IMS_ ≈ 0.018 from Eq. (4.3) shows that less than 2% of synthetic records exactly matched any real record, and those matches corresponded to highly generic profiles (e.g., healthy controls with entirely normal assessments) that were not unique within the real cohort. This pattern is consistent with low direct disclosure risk.

Second, a distance-to-closest-record (DCR) analysis on a flattened mixed-type representation compared train-to-train vs train-to-synthetic proximity ratios using Gower distance. The train-to-synthetic proximity distribution was not shifted toward smaller distances relative to the train-to-train baseline (see Fig. 2.6), yielding a privacy score 𝒫 = 1, indicating that synthetic records are not systematically closer to any given real patient than expected under internal variability (see Section 4.4).

**Figure 2.6:**
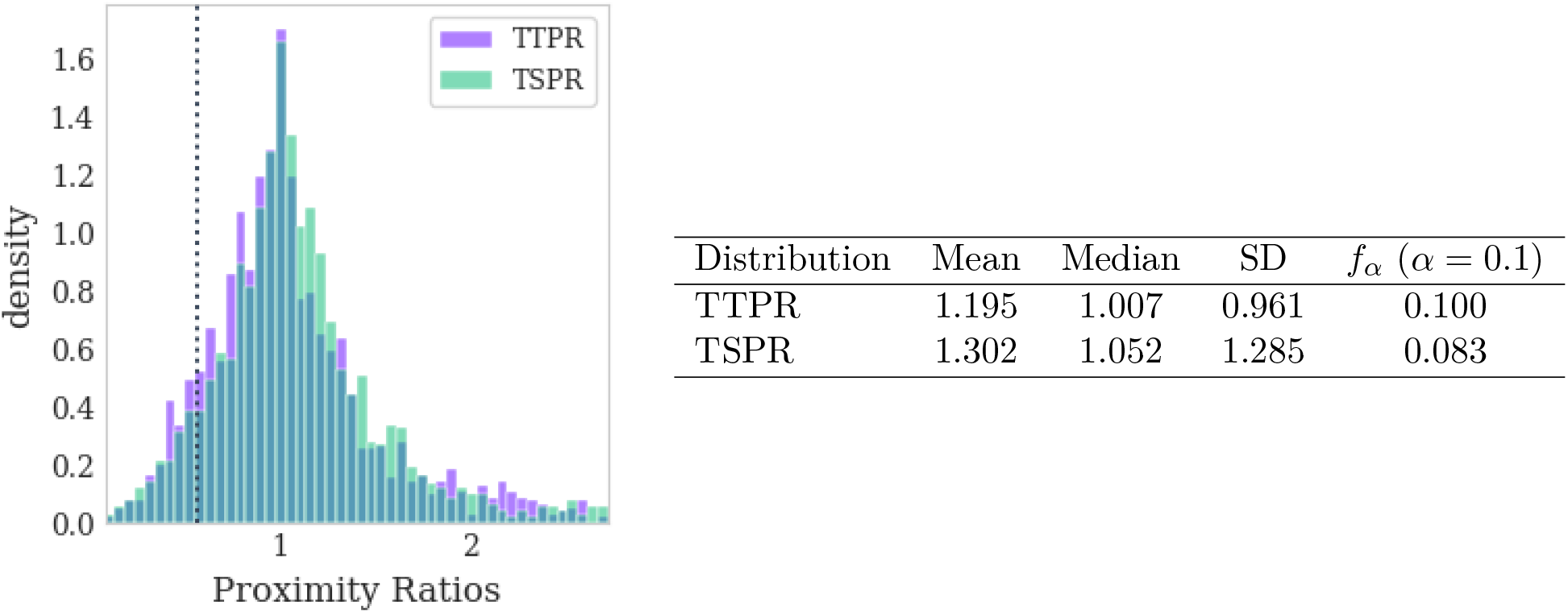
Distance-to-Closest-Record (DCR) privacy analysis on the flattened dataset. *Left:* Distributions of the Train-to-Train Proximity Ratio (TTPR, purple) and Train-to-Synthetic Proximity Ratio (TSPR, green). Dotted vertical lines indicate the *α*-quantile (*α* = 0.1) of TTPR distribution. *Right:* Summary statistics for both distributions, where *f*_*α*_ denotes the fraction of records with proximity ratio below the *α*-quantile threshold of the TTPR distribution. The Privacy Score is computed as in Eq. (4.4), obtaining 𝒫 = 1, indicating no re-identification risk.

Third, we used DOMIAS, a state-of-the-art membership inference attack tailored to tabular data, to test whether an adversary could infer whether a particular real record was included in the training set based on synthetic data. We used two conditions: Patient-level, using the root table of demographic data, and the Flattened relational data obtained by joining patient demographics with longitudinal clinical assessments and biospecimen measurements as done for DCR analysis. At both patient-level and fully flattened levels, attack AUC and accuracy were near 0.5, indistinguishable from random guessing (Table 2.4). These results fall into the low-risk category reported in prior DOMIAS benchmarks [33] and indicate minimal leakage of membership information.

**Table 2.4:** DOMIAS membership inference attack results. AUC-ROC scores near 0.5 indicate no distinguishability between training members and non-members, suggesting minimal privacy leakage.

| Evaluation Level | Accuracy | AUC-ROC |
| --- | --- | --- |
| Patient-level | $0.508 \pm 0.007$ | $0.497 \pm 0.009$ |
| Flattened relational data | $0.509 \pm 0.007$ | $0.506 \pm 0.007$ |

Taken together, these metrics demonstrate that the synthetic cohort satisfies stringent privacy requirements and can be treated as non-personal data under GDPR guidance, enabling wider sharing for research and method development while protecting individual participants.

### 2.9 Synthetic performance vs prompted GPT-5 model: fidelity comparison

A fundamental question in LLM-based synthetic data generation is whether domain-specific fine-tuning provides meaningful advantages over prompt-based generation using frontier models. To address this question directly, we present a comparison between the approach used throughout the paper (fine-tuning a smaller open-source model like Qwen3-8B-Base on the PPMI dataset), and a larger proprietary model (GPT-5-mini [34]) with in-context learning through prompting, but without any fine-tuning [27, 35, 36].

Prompt-based generation using schema descriptions and few-shot examples, without fine-tuning, produced synthetic datasets with visibly distorted univariate distributions and disrupted correlation structures (Fig. 2.7 and Fig. 2.9), in line with recent observations on text-to-tabular generation in clinical data [37]. Particularly worrisome is the absence of the inverse relationship between UPDRS motor impairment data and the imaging and alpha-synuclein CSF values, as well as the lack of association between `GM_VOLUME` and `NfL` biomarker.

**Figure 2.7:**
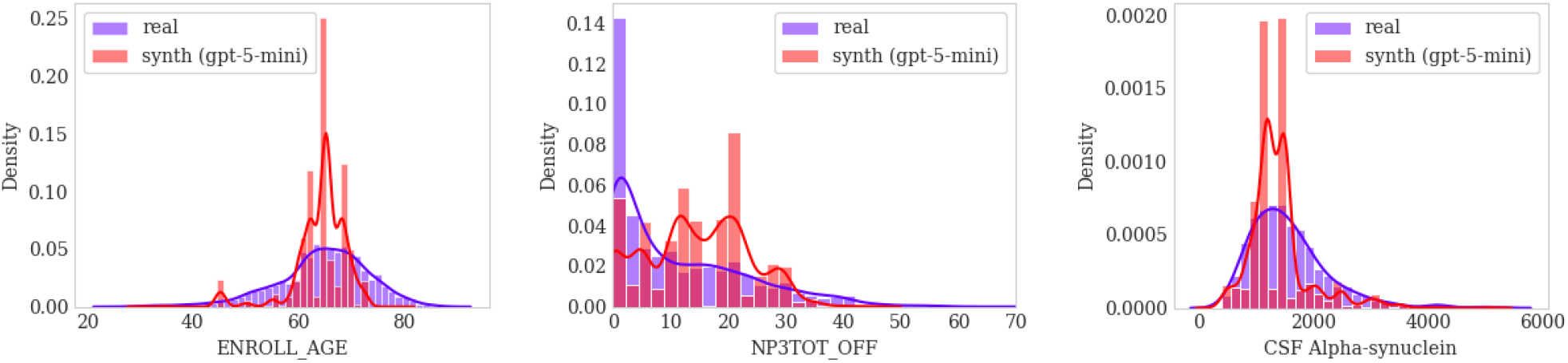
Univariate marginal distributions for selected baseline variables comparing real data (*purple*) with GPT-5-mini prompt-based synthetic data (*pink*). Histograms and kernel density estimation (KDE) curves show substantial divergence from real distributions contrasting with the close alignment achieved by fine-tuned Qwen3-8B (see Figure 2.1). The prompt-based approach struggles to capture the precise shape and support of clinical variable distributions.

**Figure 2.8:**
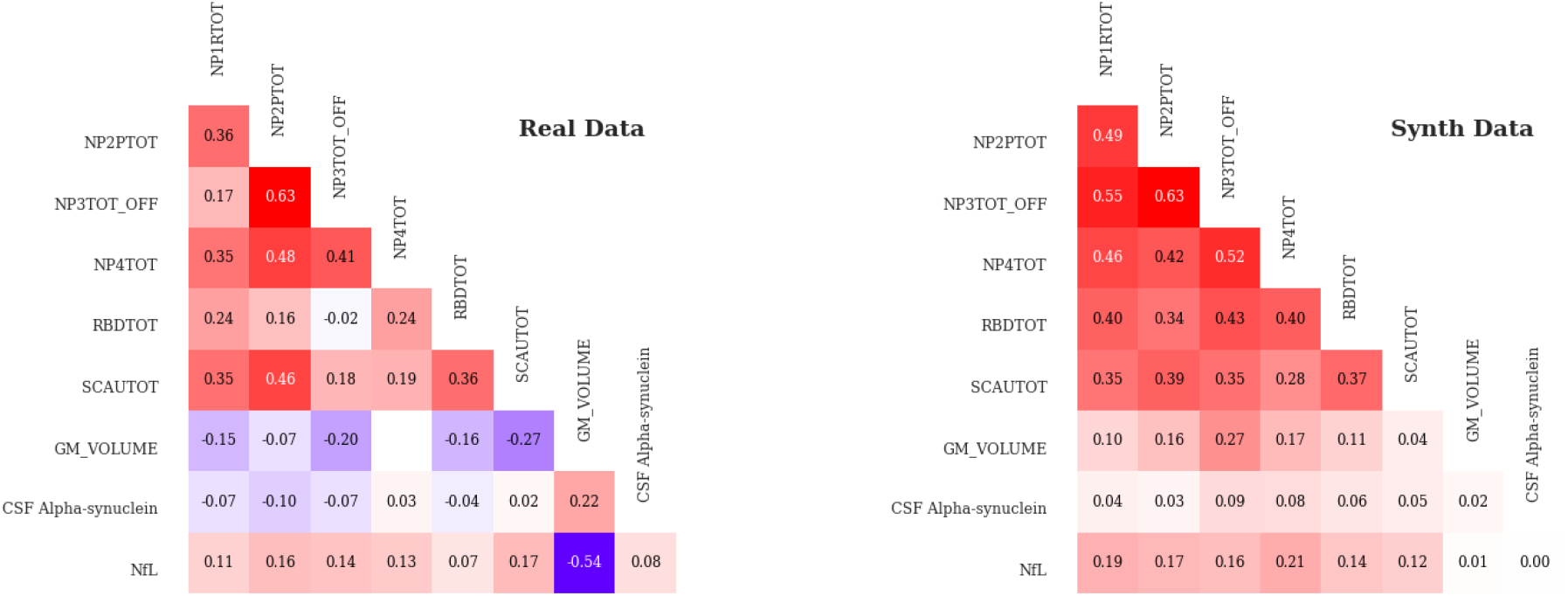
Comparison of Pearson correlation matrices at baseline between real data (*left*) and GPT-5-mini prompt-based synthetic data (*right*). The prompt-based approach shows notable deviations in correlation strength and structure compared to the alignment achieved by fine-tuned Qwen3-8B (see Figure 2.2).

**Figure 2.9:**
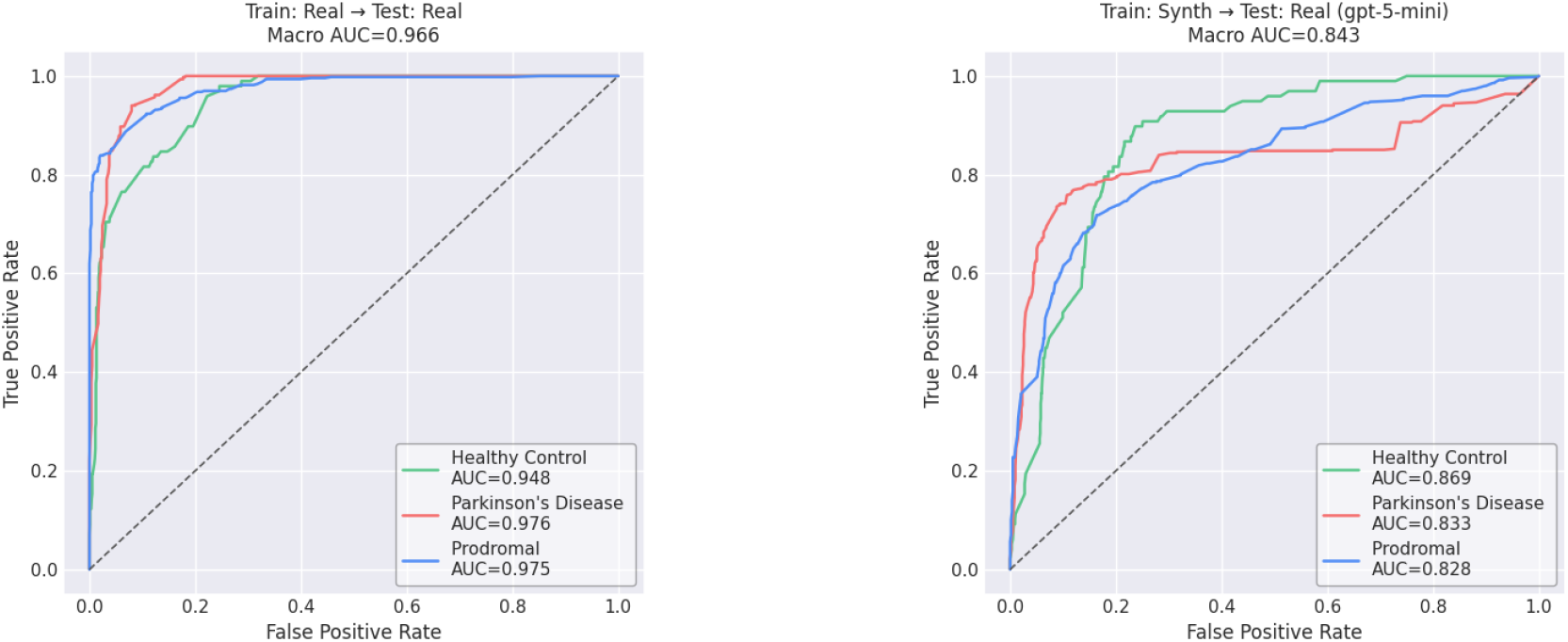
Multi-class ROC curves for Random Forest classifier. *Left:* trained and tested on real data (baseline). *Right:* trained on GPT-5-mini prompt-based synthetic data and tested on real patients. Comparing with Figure 2.4, the prompt-based approach shows degraded performance, particularly for distinguishing prodromal subjects, highlighting the utility advantage of fine-tuned synthetic data for downstream predictive modeling.

Moreover, we evaluated classification utility using the same Random Forest diagnostic model frame-work presented in Section 2.5. Fig. 2.9 shows ROC curves for the prompt-based approach, which can be directly compared with the fine-tuned results in Fig. 2.4.

These findings confirm that domain-specific fine-tuning is critical for achieving the level of distributional and mechanistic fidelity required for PD disease modeling and digital twin applications, despite the general capabilities of current frontier LLMs as GPT-5 mini.

## 3 Discussion

The present study demonstrates that domain-specific fine-tuning of a dedicated LLM generates high-fidelity virtual Parkinson’s disease patients that preserve some mechanistic disease relationships and support quantitative pharmacometric modelling of relevance for in silico trials. Three principal findings emerge: (1) LLM fine-tuning substantially outperforms prompt-based generation (e.g., GPT-5 mini) in preserving distributional fidelity and multivariate correlation structure; (2) synthetic cohorts maintain the mechanistic coupling between nigrostriatal dopaminergic pathology and motor impairment, a causally grounded axis central to PD pathophysiology; and (3) nonlinear mixed-effects disease-progression models fitted to synthetic data reproduce population parameter estimates and predictive performance obtained from real PPMI cohorts, establishing analytical utility for in silico trial simulation.

Preservation of mechanistic relationships distinguishes patients’ digital twins from generic synthetic statistically-driven datasets. Analysis of the longitudinal dopamine–motor coupling showed that synthetic data reproduced within-subject correlation and between-subject correlation consistent with real PPMI trajectories [5, 6, 11]. This quantifiable link between DaTSCAN caudate signal and MDS-UPDRS Part III OFF-state motor scores is in keeping with a causally grounded relationship established through lesion-model preclinical experiments, clinical neuroimaging, electrophysiological data, and post-mortem neuropathology [18, 19, 21, 28]. Accordingly, computational models integrating dopaminergic dynamics with large-scale brain network activity mapped with imaging or electrophysiology have further demonstrated that individual differences in striatal dopamine tone and network architecture may predict heterogeneity in symptom presentation and treatment response. The fidelity of this mechanistic axis is essential for simulating patients as digital twin applications: a generative model that fails to preserve the dopamine–motor relationship would produce biologically implausible predictions when simulating dopaminergic replacement or deep brain stimulation therapies [18,19,21]. Our results confirm that LLM fine-tuning, combined with domain-specific semantic variable descriptions and relational schema enforcement, maintains this mechanistic structure at the population level, providing the statistical and pathophysiological grounding required for patient-specific instantiation of digital twin frameworks.

Its utility was further confirmed through thorough reproduction of Ribba et al. (2024) nonlinear mixed-effects disease-progression model [20], which describes OFF-state MDS-UPDRS Part III trajectories in a clinical trial setting using a logistic growth law, with a treatment-effect expressed as LEDD that align standard-of-care dopaminergic therapeutic to the levodopa effects. Parameter estimation via the SAEM algorithm yielded population-level estimates for baseline motor severity, maximum plateau, progression rate, and treatment effect that overlapped within 95% confidence intervals between real and synthetic datasets. The sole exception was a slightly elevated maximum plateau in synthetic data, suggesting modest overestimation of long-term severity consistent with the known tendency of generative models to smooth trajectory extremes. Visual predictive checks confirmed that simulated quantiles from both fits captured the empirical distribution of motor scores across a ten-year horizon, validating preservation of mechanistic disease dynamics in the synthetic cohort. This pharmacometric replication has direct implications for in silico trial design. A recently published workflow for in silico clinical trials using nonlinear mixed-effects models demonstrated that virtual patient cohorts preserve treatment-response heterogeneity and identify biomarker-enriched populations most likely to benefit from targeted interventions [31]. The present results may extend this capability to neurodegenerative disease, where slow progression, phenotypic heterogeneity, and high placebo rates represent persistent obstacles for conventional randomised controlled trials [32].

Regulatory acceptance of in silico evidence is evolving. The recent harmonised global guideline supported by FDA, EMA and other agencies issued by ICH on guidance on Model Informed Drug Development (MIDD) qualified specific computational modelling approaches for regulatory decision-making [38]. In this framework, synthetic patients / digital twins leveraging validated pharmacometric disease-progression models can support an evidence-based strategy to accelerate the path from mechanistic hypothesis to clinical proof-of-concept, optimising clinical trial design and performance.

Stringent privacy protection is a prerequisite for broad sharing of synthetic clinical datasets. The three-layer evaluation framework [35] (IMS, DCR and DOMIAS) demonstrated compliance with GDPR Article 6 and HIPAA de-identification standards. IMS analysis revealed fewer than 2% exact matches, DCR analysis yielded a privacy score of 1.0 providing no statistical evidence of reidentification risk, and DOMIAS attack AUC-ROC values were indistinguishable from random guessing, confirming minimal membership-information leakage. Our evaluation satisfies these criteria and positions the synthetic PPMI cohort as a resource suitable for cross-institutional sharing, external model validation, and potential in silico trial simulation under GDPR-compliant data governance.

Prompt-based, general-purpose very-large LLM for health data synthetic has recently attracted some interest, but also concerns about their limitation [36]. To explore this idea and support the use of the fine-tuned, customised LLM approach, we performed a comparison between the output of our fine-tuned Qwen3-8B-Base LLM and the GPT-5-mini LLM. Despite deploying a larger frontier model with substantially greater general knowledge, GPT-5-mini prompt-based generation produced synthetic datasets with visibly distorted univariate distributions and disrupted correlation structures when compared with real data. Most consequentially, there was neither an inverse relationship between MDS-UPDRS Part III scores and DaTSCAN imaging signal, nor a positive correlation between gray matter volume loss and peripheral NfL elevation, an established biomarker of neurodegeneration. Diagnostic classification utility was correspondingly degraded, with reduced AUC values most pronounced for discrimination of prodromal subjects. Therefore, generating regulatory-grade synthetic clinical data requires not merely scale or broad pretraining, but structured learning of domain-specific constraints, relational dependencies, and temporal dynamics from in-domain examples. Fine-tuning with LoRA adapters applied to patient-level JSON objects, combined with variable-level semantic descriptions and guided JSON decoding during inference, enabled Qwen3-8B-Base to internalise these domain-specific patterns despite having substantially fewer parameters than the frontier model used for comparison. Domain-specific fine-tuning is therefore a relevant step for achieving the fidelity and mechanistic realism required for clinical research applications.

Several limitations require consideration. First, the synthetic cohort was derived exclusively from PPMI, which enrolls predominantly Caucasian participants from North America and Europe at early disease stages. Generalisability to populations with different genetic backgrounds (e.g., higher GBA or LRRK2 variant frequencies), environmental exposures, or healthcare access patterns remains to be established. External validation using geographically and ethnically diverse cohorts is an essential next step. Second, while synthetic data preserved some aggregate mechanistic structure, the present analysis is far from exhaustive: at the moment, individual synthetic patients represent plausible instantiations from the learned joint distribution, not faithful replicas of specific real individuals. Further work should examine whether rare subtypes, atypical progression patterns, and the full spectrum of motor complications and non-motor burden are adequately represented. Third, this study focused primarily on clinical assessments and dopaminergic imaging; genetic and molecular data were underutilised in model training. Incorporating pathogenic variants, polygenic risk scores, and multiomic profiles would enable biologically defined patient stratification and support genotype-informed in silico trial designs. Integration of emerging biomarkers is a high-priority extension aligned with evolving regulatory biomarker qualification frameworks [8]. Fourth, the current implementation generates static cohorts at inference time. True digital twins require continuous updating as new patient data become available, enabling dynamic recalibration of model parameters and adaptive prediction refinement. Implementing Bayesian updating or online learning schemes that incorporate longitudinal observations would advance the present simulated-patient framework towards fully dynamic digital twins capable of patient-specific trajectory forecasting.

In conclusion, domain-specific fine-tuning of a large language model on relational PPMI data produces high-fidelity virtual PD patients that could provide a practical, quantifiable pathway from real patient statistics to privacy-preserving digital twins suitable for in silico trial design, supporting model-informed drug development and precision medicine in Parkinson’s disease.

## 4 Methods

### 4.1 Synthetic Data Generation

For this study, we rely on an in-house synthetic data framework that employs LLM-based synthesis tools, building upon the capabilities of pretrained Large Language Models to generate synthetic datasets that maintain the statistical structure and analytical utility of the original data while reducing the exposure of sensitive information. The LLM-based module supports fine-tuning a pretrained language model on synthetic data generation tasks using relational tabular datasets, through which the model learns distributions, correlations, and structural patterns of the source data and produces artificial records that reflect these characteristics without reproducing actual entries.

To enable LLM-based generation while preserving patient-level independence, the relational dataset is organized into a tree structure where each root node (patient) contains all associated child records, allowing the training data to be decomposed into *independently and identically distributed* (i.i.d.) samples, each representing one complete patient with their nested clinical assessments and biospecimen measurements. We define a root table, `patients`, containing individual-level demographic and static clinical information, uniquely identified by the primary key `PATNO`. The `analyses` table captures longitudinal clinical assessments with multiple rows per participant, uniquely identified by the composite key (`PATNO, EVENT_ID`). The `biospec` table stores biospecimen measurements linked to `analyses` through the same composite key. A pictorial representation is depicted in Fig. 4.1; the full parameter list is in Table 4.1.

**Figure 4.1:**
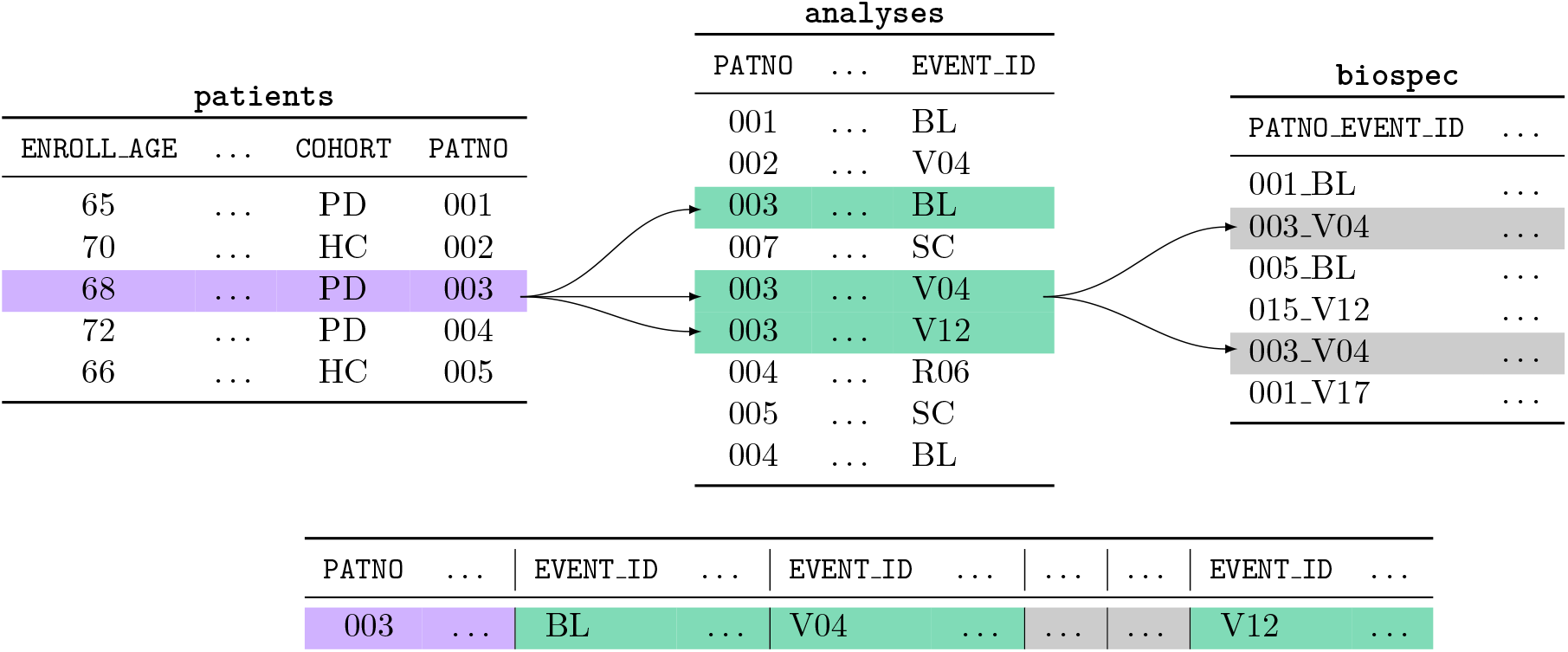
Schematic representation (*top*) of the tree-structured relational dataset for LLM-based synthetic data generation. One highlighted patient row (purple) links to three child rows in `analyses` (green), two of which link to `biospec` rows (grey); sample (*bottom*) corresponding to one root-level participant.

**Table 4.1:**
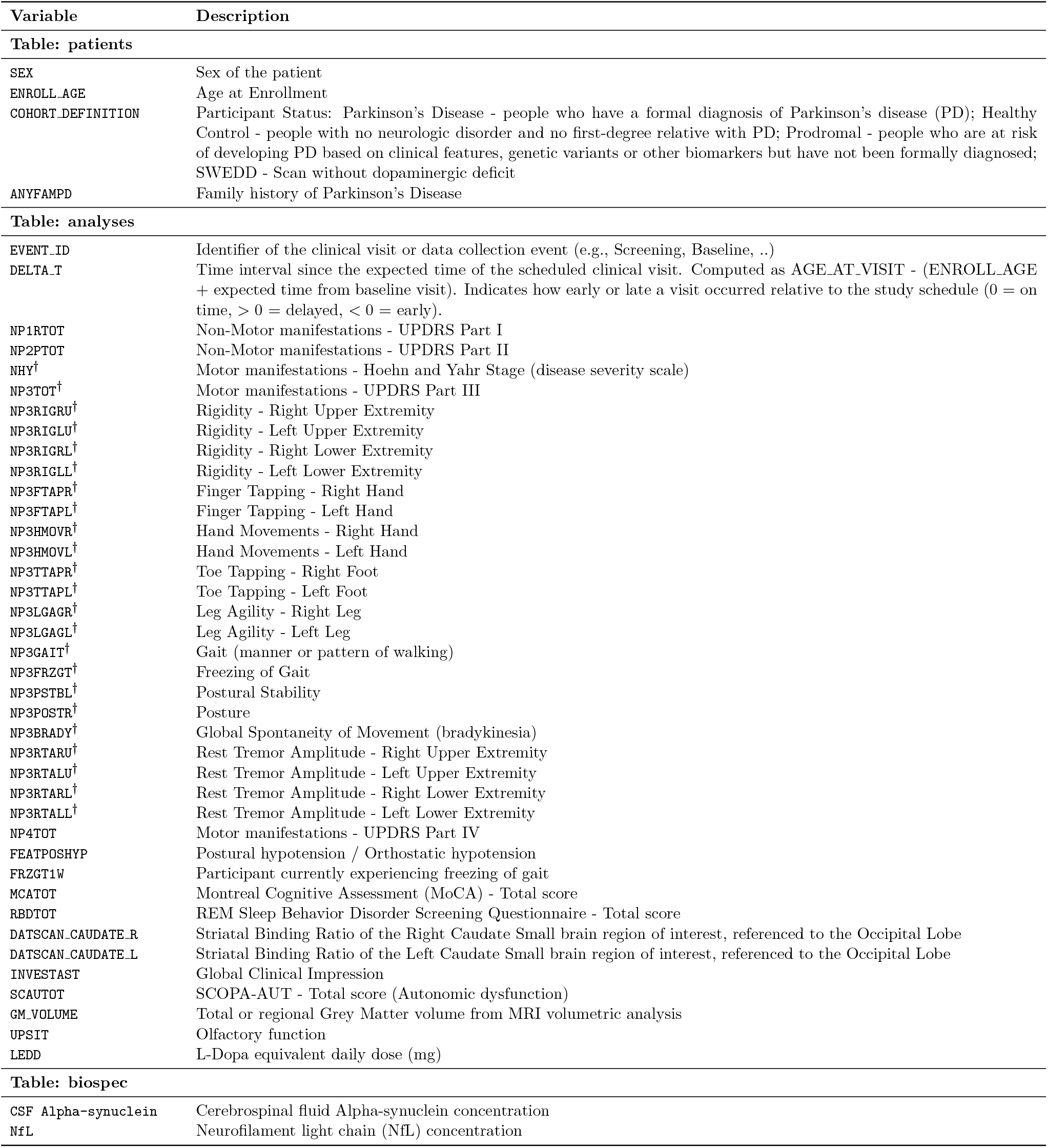
Parameters from the PPMI dataset considered for the synthesis along with their semantic descriptions provided to the LLM for each variable in the PPMI dataset. Primary and Foreign Keys are excluded (see Section 4.1). Descriptions were embedded in the training prompts to guide the model toward generating clinically plausible and internally consistent synthetic records (see [39]). ^*†*^Available in OFF-and ON-medication states.

Preprocessing transforms the relational data into independent JSON objects, each representing a single root-level participant with their nested records. JSON is adopted as the intermediate representation for its native support of hierarchical structures and compatibility with LLM structured-generation interfaces. The preprocessing step also yields a JSON schema that is passed to the inference engine to enforce structured generation, ensuring all synthetic samples conform to the expected relational layout and column-level constraints. Semantic descriptions are embedded in the prompt to guide generation (see the implementation details in [39] for a full description). Each sample is then generated from scratch as an independent root-level participant with no conditioning context.

For the task we selected Qwen3-8B-Base [26, 27], an 8.2 billion parameter language model. A base model was deliberately chosen, as base models generate text purely from pretraining patterns without bias toward structured response formats, providing a clean starting point for domain-specific adaptation. Fine-tuning used Low-Rank Adaptation (LoRA; rank *r* = 32, scaling factor *α* = 64, dropout 0.05), which freezes the original weights and trains only small adapter matrices, reducing trainable parameters to approximately 50–100 million. Training used prompt–completion pairs where the prompt encodes table names and natural-language column descriptions, and the completion is a valid JSON representation of a participant-level sample. The model was trained for up to 10 epochs with early stopping (patience of 10 evaluation periods on a 10% held-out validation set), reaching the lowest validation loss after 5 epochs. Inference was performed with vLLM, using its guided generation backend to constrain outputs to valid JSON objects conforming to the dataset schema. After inference, postprocessing routines reconstruct the generated JSON objects into relational tables, restoring primary and foreign key relationships. Full configuration and implementation details are available in the project repository [39].

### 4.2 Fidelity Assessment

Univariate fidelity was evaluated at baseline using complementary metrics applied according to variable type. For continuous variables, the Kolmogorov–Smirnov (KS) test assessed whether real and synthetic samples come from the same distribution (null hypothesis; *p >* 0.05 indicates no significant difference). For categorical variables, Kullback–Leibler (KL) divergence measured the divergence between the probability distributions of real and synthetic category proportions. Jensen–Shannon divergence (JSD) provided a symmetric, bounded information-theoretic similarity measure applicable to both continuous and discrete distributions. Multivariate mixed-type similarity was quantified using three Gower distance–based quantities: *MISS*_*R*_, the mean intra-sample similarity between real records and a randomly permuted version of the same dataset; *MISS*_*S*_, the analogous baseline computed for synthetic data; and Mean Cross-Sample Similarity (*MCSS*), the cross-sample similarity between real and synthetic records. *MISS*_*R*_ and *MISS*_*S*_ serve as within-distribution baselines representing the similarity expected between unrelated samples drawn from the same distribution.

Bivariate fidelity was evaluated through Pearson correlation matrices computed at baseline. For longitudinal variables, repeated-measures correlation (rmcorr) [30] was used to estimate within-patient associations across visits, accounting for between-individual baseline differences. Analyses were restricted to the PD cohort.

Clinical plausibility was assessed by two neurologists who independently reviewed flash cards for 20 real and 20 synthetic cases, each containing 36 demographic, clinical, imaging, and biomarker parameters. Raters provided a diagnostic judgment (PD vs. non-PD) and a plausibility rating (typical vs. implausible profile). A subset of profiles was deliberately modified as an internal validity check. Inter-rater agreement was quantified via Cohen’s *κ*, and differences between real and synthetic rating distributions were tested with Fisher’s exact test.

### 4.3 Utility Assessment

**Predictive utility** was evaluated by training a multiclass Random Forest classifier to distinguish PD, prodromal, and healthy control participants using only baseline demographics and clinical scores. Two scenarios were compared: training and testing on real data (Real → Real) as a benchmark, and training on synthetic data evaluated on held-out real patients (Synth → Real) as the operational test. Both used identical pipelines with one-hot encoding and a stratified 75%/25% split; performance was measured by class-specific AUC-ROC.

**Analytical utility** was tested by reproducing the nonlinear mixed-effects (NLME) disease progression model of Ribba et al. [20], which describes OFF-state MDS-UPDRS part III (NP3TOT OFF) trajectories via a logistic growth law combined with a treatment effect term:

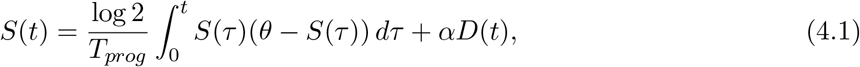

where *S*(*t*) denotes the clinical score at time *t, S*_0_ its baseline value, and *θ* its maximal plateau level. The first term captures natural disease progression through a logistic growth law parameterized by the progression rate *T*_*prog*_; the second term quantifies the impact of symptomatic treatment, with *α* representing the treatment effect (negative values indicate symptom alleviation) and *D*(*t*) the treatment exposure, operationalized as `LEDD` normalized at the individual level by its per-patient median value.

The model was implemented as a nonlinear mixed-effects system using the nlmixr2 package in R [40]. Parameter estimation employed the Stochastic Approximation Expectation Maximization (SAEM) algorithm, which is well suited for complex nonlinear models with multiple random effects. Between-subject variability in the three structural progression parameters (*S*_0_, *T*_*prog*_, *θ*) was modeled using log-normal distributions to enforce positivity, while the treatment effect *α* was modeled with a normal distribution to allow both positive and negative values. The model was fitted independently to real PPMI data and to synthetic data using identical patient selection criteria and preprocessing steps, enabling direct comparison of population-level parameter estimates and their distributions.

Model adequacy was assessed through Visual Predictive Checks (VPC), a simulation-based diagnostic that evaluates whether the fitted model reproduces the observed data distribution. For each dataset, 100 Monte Carlo replicates were generated by simulating from the fitted model with full parameter uncertainty. At each time point, prediction intervals for the 5th, 50th, and 95th percentiles of the simulated score distribution were computed and overlaid on the observed data percentiles, providing a quantitative assessment of model fit across the full follow-up horizon.

**Mechanistic utility** was assessed by analyzing the joint longitudinal relationship between the values of `NP3TOT_OFF` and `DATSCAN_CAUDATE_L` in the PD cohort over the first four years since diagnosis. For each variable separately, a Linear Mixed Model (LMM) with a fixed effect of time and a patientspecific random intercept was fitted under REML:

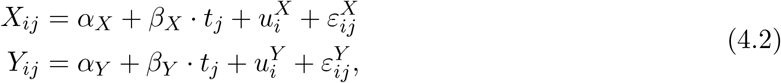

where *i* indexes patients, *j* indexes visits, *α*_*X/Y*_ is the population intercept, *β*_*X/Y*_ the population slope (units/year), 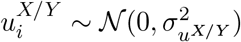 the patient-specific random intercept, and 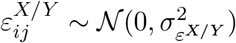 the residual error. The random-intercept-only specification was chosen to allow estimation of *ρ*_between_ from the random intercepts while keeping the within-subject slope common across patients, as required by the repeated-measures correlation framework [30].

The bivariate correlation was decomposed into two complementary components following [41, 42]. The *within-subject* component *ρ*_within_ was estimated via repeated-measures correlation on mean-centered residuals 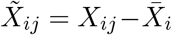 and 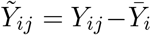 capturing the degree to which the two variables covary longitudinally *within* the same patient across visits, independently of between-patient differences in baseline levels. The *between-subject* component *ρ*_between_ was estimated as the Pearson correlation between the random intercepts 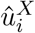 and 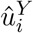 extracted from the two LMMs, capturing whether patients with higher baseline motor burden also tend to have lower dopaminergic signal at enrollment. This decomposition is important because the two components reflect distinct aspects of the dopamine–motor axis: the within-subject component captures the longitudinal coupling of dopaminergic loss and motor worsening as the disease progresses within an individual, while the between-subject component reflects the extent to which initial nigrostriatal deficit predicts initial motor severity across patients.

Patients were required to have at least three valid visits with non-missing values in both variables, yielding *n*_real_ = 329 and *n*_synth_ = 135. The smaller synthetic sample reflects the lower rate at which the generator produces records satisfying all constraints jointly, and may be underpowered to detect the weaker between-subject component. Indeed, a negative but non-significant *ρ*_between_ in a small sample is ambiguous: it could mean either that the generator did not encode the signal, or that it did but the sample is too small to detect it with adequate power. The oversampled cohort resolves this ambiguity by drawing further independent samples (*n*_over_ = 647) from the same trained model without retraining. This oversampling does not introduce new information about the real population: it only reduces finite-sample variance in characterizing the generator’s own learned distribution, thereby increasing statistical power to detect effects already encoded in the model. Bootstrap confidence intervals for *ρ*_between_ used 1000 resamples.

### 4.4 Privacy Evaluation

Privacy was assessed through three hierarchical metrics addressing progressively sophisticated disclosure risks [32, 35].

#### Identical Match Share

(IMS) quantified the risk of direct disclosure as the fraction of synthetic records exactly matching any real record across all fields:

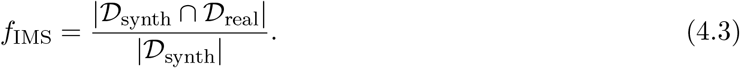

Identified matches were inspected manually to assess whether they corresponded to unique or generic profiles.

#### Distance-to-Closest-Record

(DCR) analysis quantified re-identification risk on a flattened wide-format representation of the relational dataset, obtained by pivoting all temporal measurements across visits into a single row per patient. Pairwise distances were computed using the Gower metric, which handles mixed data types by combining normalized distances for continuous variables with matching coefficients for categorical variables. The real dataset was split into two subsets *R*_1_ and *R*_2_. For each record in *R*_1_, the Train-to-Train Proximity Ratio (TTPR) was defined as the distance to its nearest neighbor in *R*_2_ divided by its distance to its second nearest neighbor in *R*_1_. Replacing *R*_2_ with the synthetic dataset *S* yields the Train-to-Synthetic Proximity Ratio (TSPR). Let *f*_*α*_(*X*) denote the fraction of records in distribution *X* whose proximity ratio falls below the *α*-quantile of the TTPR distribution (with *α* = 0.1). The Privacy Score is then defined as:

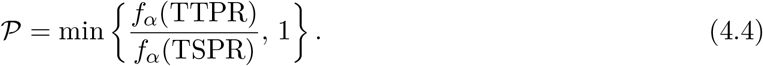

A value of 𝒫 = 1 indicates that synthetic records are not statistically closer to real records than two unrelated subsets of the real data would be to each other, implying no excess re-identification risk.

#### Membership Inference Attack

(MIA) susceptibility was evaluated using DOMIAS [33], a density ratio–based attack for tabular data. For each candidate record **x**, the attack score is:

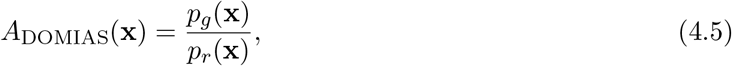

where *p*_*g*_ and *p*_*r*_ are the densities estimated from the synthetic dataset and the real training data, respectively. Records with high *A*_DOMIAS_ are predicted as training members. A balanced evaluation set (50% members, 50% non-members) was constructed by subsampling training records to match the held-out test set size; classification used the median attack score as threshold. Attack performance was measured by accuracy and AUC-ROC. Attacks were run at the patient level using Gaussian KDE for density estimation, and on the fully flattened representation using *k*-NN density estimation with *k* = 100 to handle the 3660-dimensional feature space [43,44], repeated over 10 independent runs with different random subsamples.

### 4.5 Comparison with Prompt-Based Generation

To quantify the contribution of domain-specific fine-tuning, we compared the fine-tuned Qwen3-8B-Base against GPT-5-mini [34] used with prompt-based generation only, without any fine-tuning on PPMI data. GPT-5-mini synthetic data were generated via the OpenAI batch API using the same variable semantic descriptions and relational schema, with three real patient records as few-shot examples per prompt. Fidelity was evaluated using the same univariate metrics and Pearson correlation matrix comparison described in Section 4.2. Predictive utility was assessed with the identical Random Forest pipeline, training on GPT-5-mini synthetic data and evaluating on held-out real patients.

## Data availability

Data used in the preparation of this article was obtained on 2024-06-18 from the Parkinson’s Progression Markers Initiative (PPMI) database (https://www.ppmi-info.org/access-data-specimens/download-data), RRID:SCR 006431. For up-to-date information on the study, visit www.ppmi-info.org. PPMI data are available to qualified investigators upon registration and approval, according to study-specific data use agreements.

The synthetic datasets generated in this work are derived from PPMI data. Code for LLM-based synthetic data generation, fine-tuning, and structured inference is publicly available at https://github.com/aindo-com/ppmi-llm [39], together with configuration files and full technical documentation of the implementation. No individual-level identifiable patient data are shared. The synthetic dataset is available upon reasonable request.

## Acknowledgements

We thank the Michael J. Fox Foundation and the Parkinson’s Progression Markers Initiative (PPMI) investigators and participants for providing access to high-quality longitudinal data. PPMI – a public-private partnership – is funded by the Michael J. Fox Foundation for Parkinson’s Research and funding partners, including AbbVie, Alamar Biosciences, Aligning Science Across Parkinson’s (ASAP), Arrowhead Pharma, Arvinas, AskBio, BIAL, BioArctic, Biohaven, BlueRock Therapeutics, Bristol Myers Squibb, Calico Labs, Capsida Biotherapeutics, Critical Path Institute, DaCapo Brainscience, Denali, Edmond J. Safra Foundation, Eli Lilly, Gain Therapeutics, GE Healthcare, Genentech, GSK, Insitro, Johnson & Johnson Innovative Medicine, Lundbeck, Merck, Neumora, Neuron23, Novarti, Olink, Regeneron, Roche, Sanofi, Tenvie, UCB, Vanqua Bio, Voyager Therapeutics, The Weston Family Foundation. We acknowledge the funding from a research grant of the International Foundation of Artificial Intelligence and Big Data for Human Development, Bologna, Italy. This work was also partially funded by the European Union under grant agreement No. 101218531 (SydAi).

## Author contributions

E.M.P., O.P., M.B., S.S. and M.A.M. conceived the study, defined the algorithmic and data science methodological aspect aligned with the clinical questions, and led manuscript drafting. L.L.L. and A.C. designed and implemented the LLM-based synthetic data framework and conducted generation. O.P. conducted the fidelity, utility and privacy analyses and executed disease progression and dopamine–motor modeling. M.A.M., I.C., G.C.B., M.V., C.C., E.M.P., A.B. and N.S. curated clinical data and performed neurologist evaluations. E.M.P. and J.M.F. coordinated the project, integrated clinical and modeling results, and helped in the finalization of the manuscript. All authors reviewed and approved the final version.

